# Quality of life, symptom burden and associated factors among lung cancer patients: baseline study of three public hospitals across sub- Saharan Africa

**DOI:** 10.1101/2024.04.29.24306530

**Authors:** Mpho Ratshikana, Kija Malale, Lawrence Atundo Murunga, Abdul-Rauf Sayed, Oluwatosin Ayeni, Themba G Ginindza, Xolisile Dlamini, Naftali Busakhala, Rofhiwa Mathiba, Charmaine Blanchard, Merika Tsitsi, Sabelo Msomi, Nestory Masalu, Herry O. Dhudha, Paul Ruff, Anita Graham, Daniel Osei-Fofie

## Abstract

**Background:** Cancer patients in Sub-Saharan Africa (SSA) are diagnosed late partly due to community lack of knowledge about the disease, social and cultural factors, health system challenges, and inadequate health care worker knowledge. These delays in diagnosis as well as inadequate treatment options contribute to the high mortality from lung cancer in SSA. Quality of life (QoL) is an important outcome measure for cancer patients undergoing treatment.

**Objective:** To describe the quality of life among lung cancer patients in three teaching hospitals in SSA.

**Methods:** This is a prospective cross-sectional study of lung cancer patients at three teaching hospitals in Sub-Saharan Africa (SSA-Kenya (BMC), Tanzania (MTRH) and South Africa (The Lung Laboratory Research and Intervention Unit Helen Joseph Hospital). Trained interviewers collected data on demographics, clinical information and performance status using the Eastern Cooperative Oncological Group Performance Scale (ECOG-PS). Patients’ QoL was assessed using the 30-item European Organization for Research and Treatment of Cancer Quality of Life Questionnaire Core 30 (EORTC QLQ-C30).

**Results:** A total of 210 lung cancer patients consented and were enrolled across the three sites. Global Health Status in this cohort is low, the median score was 41.7 (range: 0-100) and differed between sites. Wits Core patients had higher social functioning, while BMC and MTRH had higher financial difficulty scores. Poor ECOG-PS score (3-4) was associated with poorer Global QoL (GqoL) score (aOR = 2.9; 95% CI: 1.4 - 5.9), and patients with higher symptom burden had poorer GQoL.

**Conclusion:** The QoL among lung cancer patients in the three sites is low. Poor QOL in the study is associated with level of education, performance status, fatigue, pain, dyspnoea, insomnia, loss of appetite and constipation.

## Introduction

Globocan estimates that over 19 million new cancer cases were reported worldwide in 2020, with half resulting in death. Lung cancer is the 2^nd^ most common cancer worldwide among both males and females at 11.4%, but remains the most common cause of death at 18%^1^. Although lung cancer incidence is lower in Africa, compared to more developed countries, it ranks in the top 5 common cancers in both genders and is the 4^th^ most common cause of death due to cancer. Southern Africa has the highest incidence of lung cancer on the continent^1^. Cancer patients in Sub-Saharan Africa (SSA) are diagnosed late partly due to community lack of knowledge about the disease^2^. In addition, social and cultural factors, health system challenges, and inadequate health care worker knowledge, further compromise timely diagnosis and optimal cancer care^3 4 5^ . These delays in diagnosis as well as inadequate treatment options contribute to the high mortality from lung cancer in SSA^6^. Common symptoms of lung cancer include fatigue, loss of appetite, dyspnoea, cough, pain, and haemoptysis. However, in SSA, common presenting symptoms include neurological deficits due to brain metastases, pleural effusion, spinal cord compression and superior vena cava obstruction syndrome, due to late presentation and diagnosis^7 8^. Treatment is limited to palliative chemotherapy, radiation and other interventions aimed at improving the quality of life (QoL).

Quality of life is an important outcome measure for cancer patients undergoing treatment^9 10 11^. Lung cancer diagnosis is associated with high morbidity which can be optimized by instituting measures to improve QoL. Deterioration in the health status, physical, social, spiritual and emotional wellbeing among advanced lung cancer patients is well documented, with reports of an association between the QoL and common symptoms, clinical characteristics, and sociodemographic factors ^12 13^. Assessing and improving the QoL among these patients is important, because health related QoL is associated with health outcomes and survival^9 14^. Cancer treatments including medical (chemotherapy, targeted therapies and immunotherapy) and radiation therapy and palliative care are associated with improved lung function, physical function, symptoms, and mood, leading to better QoL and survival^10 15^. Research on the symptom burden and QoL of lung cancer patients originates mostly from high income countries and is limited in SSA.

The aim of this study is to describe the common symptoms and QoL of lung cancer patients treated at three different hospitals in SSA: Cancer and Chronic Disease Centre (Kenya), Helen Joseph Hospital (South Africa) and Bugando Medical Centre (Tanzania), and to investigate the association with demographic and clinical characteristics. Our hypothesis is that the lung cancer patients in SSA will have high symptom burden, poor QoL, and that there will be an association between the demographic factors, clinical characteristics and QoL of lung cancer patients.

## Methods

### Setting

The study was conducted at three teaching hospitals in three countries, two Low to Middle Income Countries (LMIC); Kenya and Tanzania, and South Africa, an Upper Middle-Income Country (UMIC). In Kenya the Chandaria Cancer and Chronic Diseases Centre (CCCDC) is based at the Moi Teaching and Referral Hospital (MTRH) in Eldoret. MTRH, an affiliate of the Moi University School of Medicine is the second-largest National Teaching and Referral Hospital (level 6 Public Hospital) after the Kenyatta National Hospital. The CCCDC provides comprehensive cancer care services to patients mainly from Western Kenya, which includes North-Rift, Western and Nyanza regions with over 15 counties. It also serves populations from neighbouring East African countries such as, Tanzania, Rwanda, Uganda, and South Sudan. The Lung Laboratory Research and Intervention Unit Helen Joseph Hospital (Lung Lab) is a specialized respiratory unit providing comprehensive lung cancer services to patients in Johannesburg, South Africa. It is based at Helen Joseph Hospital, a teaching hospital affiliated with the University of Witwatersrand (Wits). Bugando Medical Centre (BMC) is a consultant, teaching and referral hospital for the Lake and Western zone of the United Republic of Tanzania (URT) linked to the Catholic University of Health and Allied Sciences in Mwanza, Tanzania. The hospital’s Oncology department is the only cancer centre in the Lake and Western zone of URT that provides comprehensive cancer care with outreach services to urban and rural areas.

### Design and participants

This is a cross-sectional study of lung cancer patients at three teaching hospitals in Sub-Saharan Africa (SSA). Data was collected from 1^st^ June 2018 to 31 December 2020. Inclusion criteria were patients 18 years or older, confirmed primary lung cancer diagnosis, and physically and mentally able to participate in the study. At all sites, once diagnosis was confirmed and before treatment began, patients were invited to participate in the study. Formal consent was obtained, and those who consented were enrolled and sociodemographic and clinical information was collected. Thereafter, the QLQ C30 questionnaire was administered in English and where needed translated into the vernacular.

### Study measures

Trained nurses/interviewers conducted interviews to obtain demographics (age, gender, marital status, education level, employment status, smoking, mining history), and clinical information (histology, clinical staging, ECOG, weight loss, active or current TB and other comorbidities). The Eastern Cooperative Oncological Group Performance Scale (ECOG) was used to determine the performance status of patients^16^. Patients’ QoL was assessed using the 30-item European Organization for Research and Treatment of Cancer Quality of Life Questionnaire Core 30 (EORTC QLQ-C30)^17^.The questionnaire contains five multi-item function scales (physical, role, social, emotional and cognitive functions), three multi-item symptom scales (fatigue, pain, nausea), and five single items (dyspnoea, insomnia, appetite loss, constipation, diarrhoea). The final item evaluates patient perceived financial difficulties. Each item has four possible responses alternatives: (1) “not at all”, (2) “a little”, (3) “quite a bit”, and (4) “very much”. The items are grouped to arrive at a global health status/QoL score. The responses to the scale items refer to “last week,” with the exception of the patient’s physical performance scale, where the timeframe is the present. The scores of all the Health Related QoL (HRQoL) items were calculated in accordance with the EORTC QLQ-C 30 scoring manual^11^.The sum of items in each category is added and the total divided by the number of questions in the category. A linear transformation is then undertaken to convert this to a percentage scale. All the scales and single-item measures range from 0 to 100. Higher scores on the functional and quality of life scales translated to better HRQoL, whereas higher scores on the symptom scales translated to a higher level of symptoms/problems.

### Data analysis

Study data were collected and managed using REDCap electronic data capture tools hosted at MTRH, (CCCDC building) and Wits. Patient’s demographic, clinical and health related QoL data were extracted from the REDCap database and exported to Stata 13.1 for statistical analysis^18 19^. Categorical variables are presented as frequency tables, and continuous variables are presented as descriptive measures, expressed as median and range. Non-parametric Kruskal Wallis and Wilcoxon Rank-Sum test was used to assess the differences in global health status, functional and symptom scales. The association between Global Quality of Life (GQoL) and selected socio-demographic and clinical variables (age, gender, education, occupation, smoking, MLCCP site, clinical staging, ECOG performance, comorbidity) was assessed using bivariate and multivariate logistic regression analysis. The dependent variable (GQoL) was categorized as a binary variable; a score greater than or equal to 50 was defined as “above average GQoL”. Odds ratios (OR) were used to test the association between binary variables and 95% confidence intervals (CI) that did not span unity were considered as thresholds of statistical significance. Adjusted odds ratios (aOR) were used in multivariate analysis.

### Ethics approval

Ethical approval was obtained from the University of Witwatersrand Human Research Ethics Committee (Medical) (Ref: M180436), MTRH Institutional Research and Ethics Committee IREC (0004048), BMC/CUHAS Ethics & Review Committee (Certificate number CREC/278/2018) and National Institute for Medical Research (Certificate number MR/53/100/598).

### Socio-demographic characteristics

A total of 210 lung cancer patients consented and were enrolled across the three sites: 35 at BMC, 32 at MTRH and 143 at the Lung Lab. Table 1 depicts the socio-demographic characteristics of the participants. Most patients were male (62.9%), unemployed or retired (80%), with a mean (SD) age of 61.5 (11.9) years. There were significant differences in the demographic characteristics between the patients in the three MLCCP sites. More Lung Lab patients were males (69.2%) compared to the patients in BMC (51.4%) and MTRH (46.9%) (p=0.019). Over two-thirds of the lung cancer patients at The Lung Laboratory Research and Intervention Unit Helen Joseph Hospital had high school or higher level of education compared to BMC (22.9%) and MTRH (40.6%) (p<0.001). A significant proportion of the Lung Lab patients (76.9%) were current or ex-smokers compared to patients at BMC (22.9%) and MTRH (18.7%) (p<0.001).

**Table 1.** Socio-demographic characteristics of lung cancer patients.

| Table 1 Socio-demographic characteristics of lung cancer patients |  |  |  |  |  |  |  |  |  |
| --- | --- | --- | --- | --- | --- | --- | --- | --- | --- |
|  | MLCCP site |  |  |  |  |  |  |  |  |
|  | BMC |  | MTRH |  | Lung Lab |  | Total |  | p-value |
| Socio-demographic variables | N=35 |  | N=32 |  | N=143 |  | N=210 |  |  |
|  | n | % | n | % | n | % | n | % |  |
| Age-group |  |  |  |  |  |  |  |  |  |
| <40 | 4 | 11.4 | 1 | 3.1 | 5 | 3.5 | 10 | 4.8 |  |
| 40-49 | 4 | 11.4 | 5 | 15.6 | 10 | 7.0 | 19 | 9.1 |  |
| 50-59 | 10 | 28.6 | 10 | 31.3 | 35 | 24.5 | 55 | 26.2 |  |
| 60-69 | 11 | 31.4 | 6 | 18.8 | 56 | 39.2 | 73 | 34.8 |  |
| 70+ | 6 | 17.1 | 10 | 31.3 | 37 | 25.9 | 53 | 25.2 |  |
| Mean (SD) age in years | 58 (13.4) |  | 61.5 (13.2) |  | 62.4 (11.1) |  | 61.5 (11.9) |  | 0.203 |
| Sex |  |  |  |  |  |  |  |  |  |
| Female | 17 | 48.6 | 17 | 53.1 | 44 | 30.8 | 78 | 37.1 |  |
| Male | 18 | 51.4 | 15 | 46.9 | 99 | 69.2 | 132 | 62.9 | 0.019 |
| Educational level |  |  |  |  |  |  |  |  |  |
| ≤ Primary education | 27 | 77.1 | 19 | 59.4 | 43 | 30.1 | 89 | 42.4 |  |
| ≥ High school | 8 | 22.9 | 13 | 40.6 | 100 | 69.9 | 121 | 57.6 | <0.001 |
| Marital status† |  |  |  |  |  |  |  |  |  |
| Married or in Partnership | 29 | 82.9 | 25 | 78.1 | 53 | 45.7 | 107 | 58.5 |  |
| Single/divorced/widowed | 6 | 17.1 | 7 | 21.9 | 63 | 54.3 | 76 | 41.5 | <0.001 |
| Occupation |  |  |  |  |  |  |  |  |  |
| Employed | 10 | 28.6 | 3 | 9.4 | 29 | 20.3 | 42 | 20.0 |  |
| Unemployed/retired | 25 | 71.4 | 29 | 90.6 | 114 | 79.7 | 168 | 80.0 | 0.144 |
| Smoking |  |  |  |  |  |  |  |  |  |
| Never smoked | 27 | 77.1 | 26 | 81.3 | 33 | 23.1 | 86 | 41.0 |  |
| Current or ex-smoker | 8 | 22.9 | 6 | 18.7 | 110 | 76.9 | 124 | 59.0 | <0.001 |
| Ever a miner |  |  |  |  |  |  |  |  |  |
| Yes‡ |  |  |  |  | 19 | 13.3 | 19 | 9.1 |  |
| No | 35 | 100.0 | 32 | 100.0 | 124 | 86.7 | 191 | 91.0 |  |
| † Marital status was unknown in 27 Lung Lab patients |  |  |  |  |  |  |  |  |  |
| ‡ None of the BMC and MTRH patients ever worked in a mine |  |  |  |  |  |  |  |  |  |

### Clinical characteristics

The majority of patients (90%) were diagnosed with non-small cell lung cancer (NSCLC). Most presented with late stage (III-IV) diseases; 97.3% of the NSCLC (n=188) and 86.4% of the Small Cell Lung Cancer (SCLC) (n=22). Only 5 of The Lung Laboratory Research and Intervention Unit Helen Joseph Hospital patients (3.9%) with NSCLC presented with early stage (I-II) (Table 2). The majority of patients at BMC (85.7%) presented with weight loss compared to patients at MTRH (53.1%) and the Lung Lab (67.1%) (p=0.015). Similarly, a significant proportion of BMC patients had current TB (34.3%) compared to the patients in MTHR (9.4%) and the Lung Lab (3.5%) (p<0.001). Almost all MTRH patients (96.9%) reported a near normal performance status (ECOG 0-2) compared to the patients at BMC (65.7%) and the Lung Lab (72.7%) (p=0.006). A higher proportion of the Lung Lab patients presented with one or more comorbidity (58%) compared to patients in BMC (40%) and MTRH (25%) (P<0.01).

**Table 2.** Clinical characteristics of lung cancer patients.

| Table 2 Clinical characteristics of lung cancer patients |  |  |  |  |  |  |  |  |  |
| --- | --- | --- | --- | --- | --- | --- | --- | --- | --- |
| MLCCP site |  |  |  |  |  |  |  |  |  |
|  | BMC |  | MTRH |  | Lung Lab |  | Total |  | p-value |
| Clinical variables | N=35 |  | N=32 |  | N=143 |  | N=210 |  |  |
|  | n | % | n | % | n | % | n | % |  |
| Clinical staging |  |  |  |  |  |  |  |  |  |
| NSCLC | n=27 |  | n=31 |  | n=130 |  | n=188 |  |  |
| I-II | 0 | 0 | 0 | 0 | 5 | 3.9 | 5 | 2.7 |  |
| III-IV | 27 | 100.0 | 31 | 100.0 | 125 | 96.2 | 183 | 97.3 | 0.318 |
| SCLC | n=8 |  | n=1 |  | n=13 |  | n=22 |  |  |
| Limited | 0 | 0.0 | 0 | 0.0 | 3 | 23.1 | 3 | 13.6 |  |
| Extensive | 8 | 100.0 | 1 | 100.0 | 10 | 76.9 | 19 | 86.4 | 0.300 |
| ECOG performance |  |  |  |  |  |  |  |  |  |
| 0-2 | 23 | 65.7 | 31 | 96.9 | 104 | 72.7 | 158 | 75.2 |  |
| 3-4 | 12 | 34.3 | 1 | 3.1 | 39 | 27.3 | 52 | 24.8 | 0.006 |
| Weight loss |  |  |  |  |  |  |  |  |  |
| No | 5 | 14.3 | 15 | 46.9 | 47 | 32.9 | 67 | 31.9 |  |
| Yes | 30 | 85.7 | 17 | 53.1 | 96 | 67.1 | 143 | 68.1 | 0.015 |
| Current TB |  |  |  |  |  |  |  |  |  |
| No | 23 | 65.7 | 29 | 90.6 | 138 | 96.5 | 190 | 90.5 |  |
| Yes | 12 | 34.3 | 3 | 9.4 | 5 | 3.5 | 20 | 9.5 | <0.001 |
| HIV status |  |  |  |  |  |  |  |  |  |
| HIV negative | 35 | 100.0 | 0 |  | 120 | 83.9 | 155 | 73.8 |  |
| HIV positive | 0 |  | 0 |  | 23 | 16.1 | 23 | 11.0 |  |
| Unknown † | 0 |  | 32 | 100.0 | 0 | 0.0 | 32 | 15.2 |  |
| Comorbidity |  |  |  |  |  |  |  |  |  |
| No | 21 | 60.0 | 24 | 75.0 | 60 | 42.0 | 105 | 50.0 |  |
| Yes | 14 | 40.0 | 8 | 25.0 | 83 | 58.0 | 105 | 50.0 | 0.001 |

### Quality of life assessment (EORTC QLQ-C30)

Overall, the EORTC QLQ-C30 Global Health Status (GHS) was low (Table 3); the median score was 41.7 (range 0-100). The GHS score was significantly higher among patients in MTRH (50.0) compared to BMC (33.3) and the Lung Lab (41.7) (p<0.001). Lung Lab patients had higher social functioning scores compared to patients in BMC and MTRH (p<0.001). Financial difficulty was significantly higher among BMC and MTRH patients compared to Lung Lab (p<0.001). Both Lung Lab and BMC cohorts had higher cognitive functioning scores compared to MTRH (p=0.0182). Overall, the highest symptom scores reported were pain (66.7) and fatigue (55.6). Pain and insomnia scores were significantly higher among patients in BMC, with median scores 83.3 (p=0.0028) and 66.7 (p=0.0024) respectively compared to Lung Lab and MTRH.

**Table 3:** Global health status, functional scales, and symptom scales of lung cancer patients. ^20^.

| Assessment<br>of quality-<br>of-life scale<br>(QLQ-C30) | Reference<br>N= 3,332 |  | Total<br>N=210 |  | BMC<br>N=35 |  | MTRH<br>N=32 |  | Lung Lab<br>N=143 |  | P-value |
| --- | --- | --- | --- | --- | --- | --- | --- | --- | --- | --- | --- |
|  | Median | IQR | Median | IQR | Median | IQR | Median | IQR | Median | IQR |  |
| Global<br>health<br>status-QL | 58.3 | 41.7-75 | 41.7 | 33.3 -<br>58.3 | 33.3 | 16.7 -<br>50.0 | 50.0 | 41.7 -<br>66.7 | 41.7 | 33.3 -<br>58.3 | 0.0001* |
| **Functional**<br>**scales** |  |  |  |  |  |  |  |  |  |  |  |
| Physical functioning- PF | 80 | 60-86.7 | 53.3 | 26.7 - 73.3 | 66.7 | 6.7 - 86.7 | 53.3 | 30 - 66.7 | 46.7 | 33.3 - 73.3 | 0.7156 |
| Role functioning- RF | 66.7 | 33.3-100 | 33.3 | 16.7 - 66.7 | 50.0 | 0.0 - 83.3 | 25.0 | 0 - 66.7 | 33.3 | 16.7 - 66.7 | 0.3197 |
| Emotional functioning- EF | 75 | 50-91.7 | 66.7 | 50.0 - 83.3 | 75.0 | 41.7 - 100 | 66.7 | 37.5 - 83.3 | 66.7 | 50 - 83.3 | 0.5347 |
| Cognitive functioning- CF | 83.3 | 50-100 | 66.7 | 50.0 - 83.3 | 83.3 | 50.0 - 100 | 58.3 | 25 - 83.3 | 83.3 | 50 - 83.3 | 0.0182* |
| Social functioning- SF | 83.3 | 50-100 | 33.3 | 0.0 - 66.7 | 0.0 | 0 - 0 | 33.3 | 0 - 33.3 | 50.0 | 33.3 - 83.3 | 0.0001* |
| **Symptoms scale** |  |  |  |  |  |  |  |  |  |  |  |
| Fatigue- FA | 33.3 | 22.2-55.6 | 55.6 | 33.3 - 77.8 | 55.6 | 33.3 - 100 | 55.6 | 22.2 - 66.7 | 55.6 | 44.4 - 77.8 | 0.1153 |
| Nausea and vomiting- NV | 0 | 0-16.7 | 0.0 | 0.0 - 16.7 | 16.7 | 0 - 66.7 | 0.0 | 0 - 16.7 | 0.0 | 0 - 16.7 | 0.0544 |
| Pain- PA | 16.7 | 0-50 | 66.7 | 33.3 -<br>83.3 | 83.3 | 50 -<br>100 | 66.7 | 33.3 -<br>83.3 | 50.0 | 33.3 -<br>66.7 | 0.0028* |
| Dyspnoea-DY | 33.3 | 0-66.7 | 33.3 | 33.3 -<br>66.7 | 33.3 | 0 - 100 | 66.7 | 33.3 -<br>66.7 | 33.3 | 33.3 -<br>66.7 | 0.6227 |
| Insomnia-SL | 33.3 | 0-66.7 | 33.3 | 0.0 - 66.7 | 66.7 | 33 -<br>100 | 33.3 | 0 - 66.7 | 33.3 | 0 - 66.7 | 0.0024* |
| Appetite<br>loss- AP | 0 | 0-66.7 | 33.3 | 0.0 - 66.7 | 33.3 | 0 - 100 | 33.3 | 0 - 66.7 | 33.3 | 0 - 66.7 | 0.4228 |
| Constipation-CO | 0 | 0-33.3 | 0.0 | 0.0 - 33.3 | 0.0 | 0 - 0 | 0.0 | 0 - 0 | 0.0 | 0 - 33.3 | 0.0001* |
| Diarrhoea-DI | 0 | 0-0 | 0.0 | 0 - 0 | 0.0 | 0 - 33.3 | 0.0 | 0 - 0 | 0.0 | 0 - 0 | 0.0041* |
| Financial<br>difficulties -<br>FI | 0 | 0-33.3 | 66.7 | 33.3 -<br>100 | 100.0 | 100 -<br>100 | 100.0 | 100 -<br>100 | 66.7 | 33.3 -<br>66.7 | 0.0001* |

\*Significant at $p < 0.05$ . Abbreviations: BMC (Bugando Medical Centre), MTRH (Moi Teaching and Referral Hospital), Lung Lab (Helen Joseph Hospital)

The results of the multiple logistic regression analysis using the dependent variable (GQoL) as a binary outcome (GQoL score <50 and GQoL score ≥50) are shown in Table 4. A significantly higher proportion of lung cancer patients at BMC and The Lung Lab had below average GQoL compared to patients at MTRH (aOR = 5.0; 95% CI: 1.6 - 15.6) and (aOR = 2.9; 95% CI: 1.0 - 8.0) respectively. Poor ECOG score (3-4) was associated with poorer GQoL score (aOR = 2.9; 95% CI: 1.4 - 5.9). The bivariate analysis showed a significant association between GQoL and educational level. Patients with below secondary level of education were more likely to have below average GQoL, however the multivariate analysis showed no significant associations when adjusted for all demographic variables (age, gender, education, occupation and smoking).

**Table 4:**
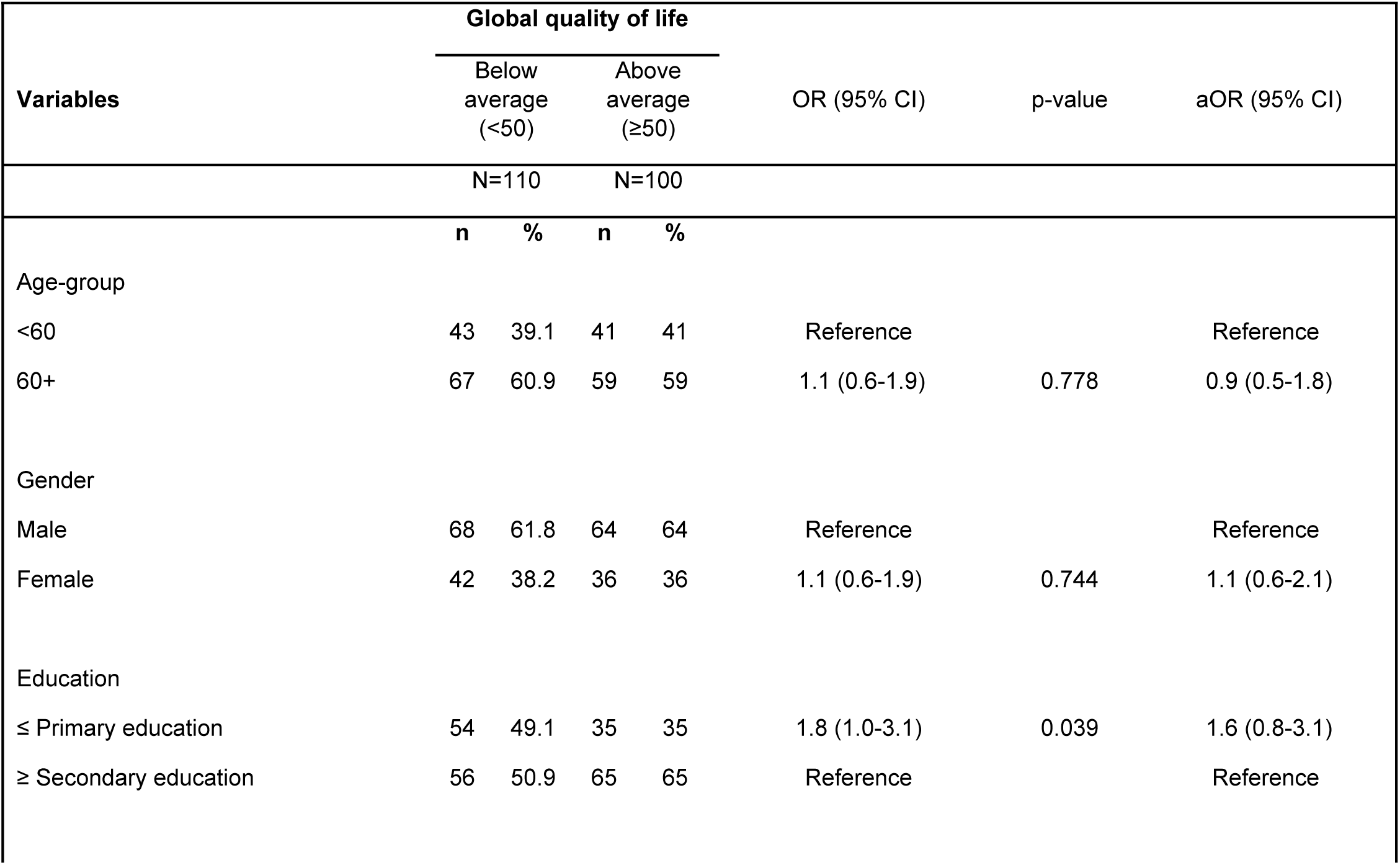
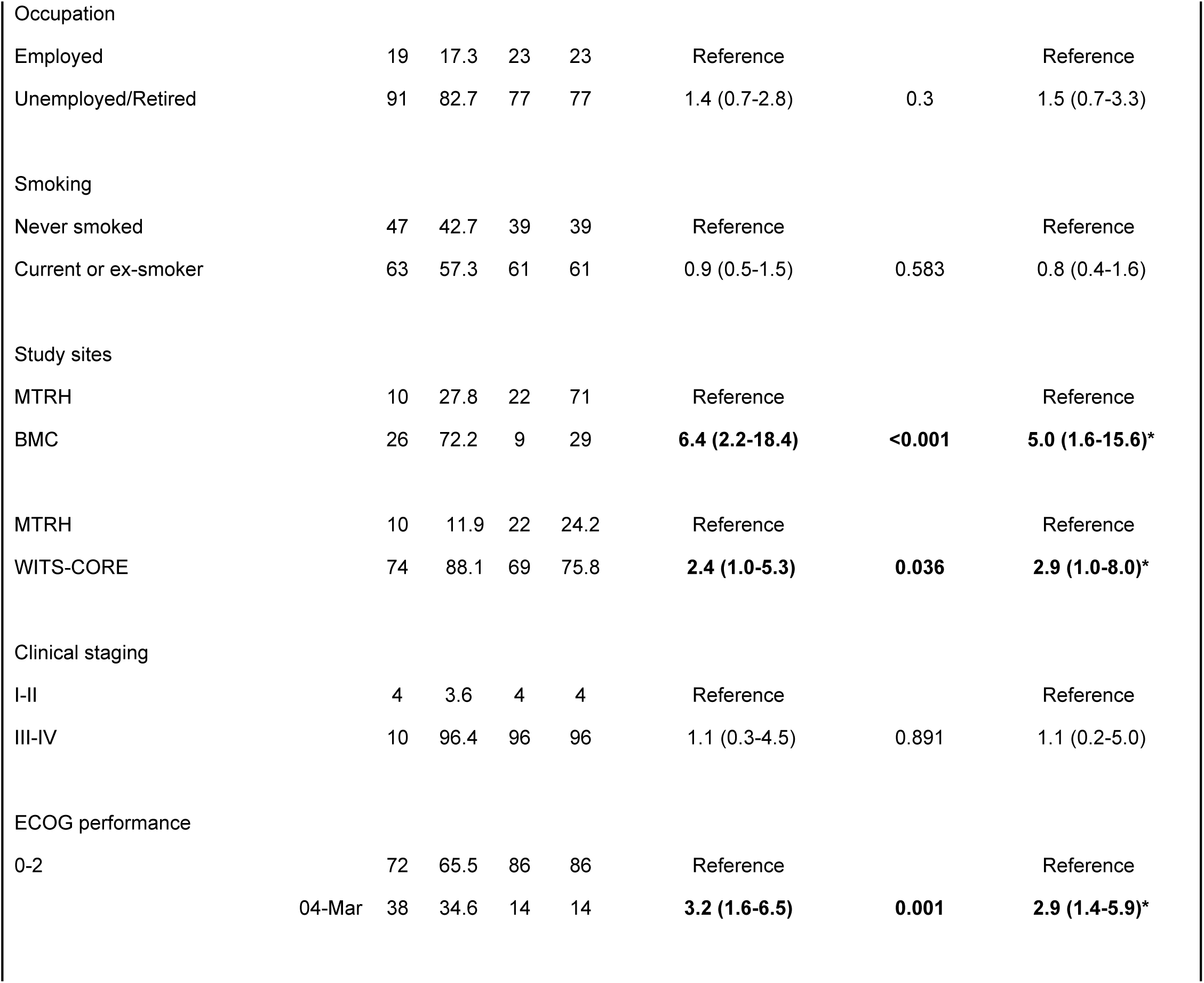
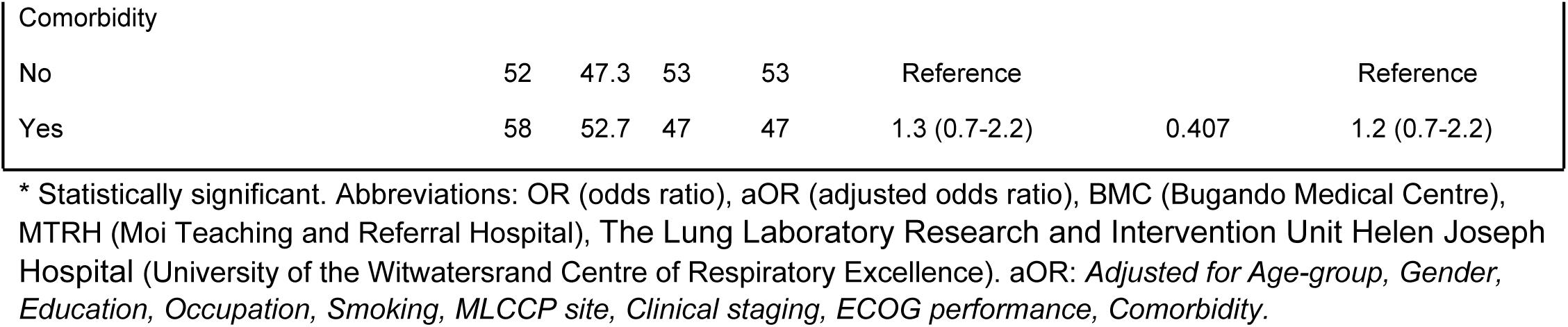
Associations between socio-demographic, clinical characteristics, and global quality of life.

The symptom scores were significantly higher among patients with below average GQoL (<50) for fatigue, pain, dyspnoea, insomnia, appetite loss and constipation when compared to above average GqoL (≥50) (Table 5). Higher symptom scores are associated with global health status.

**Table 5:**
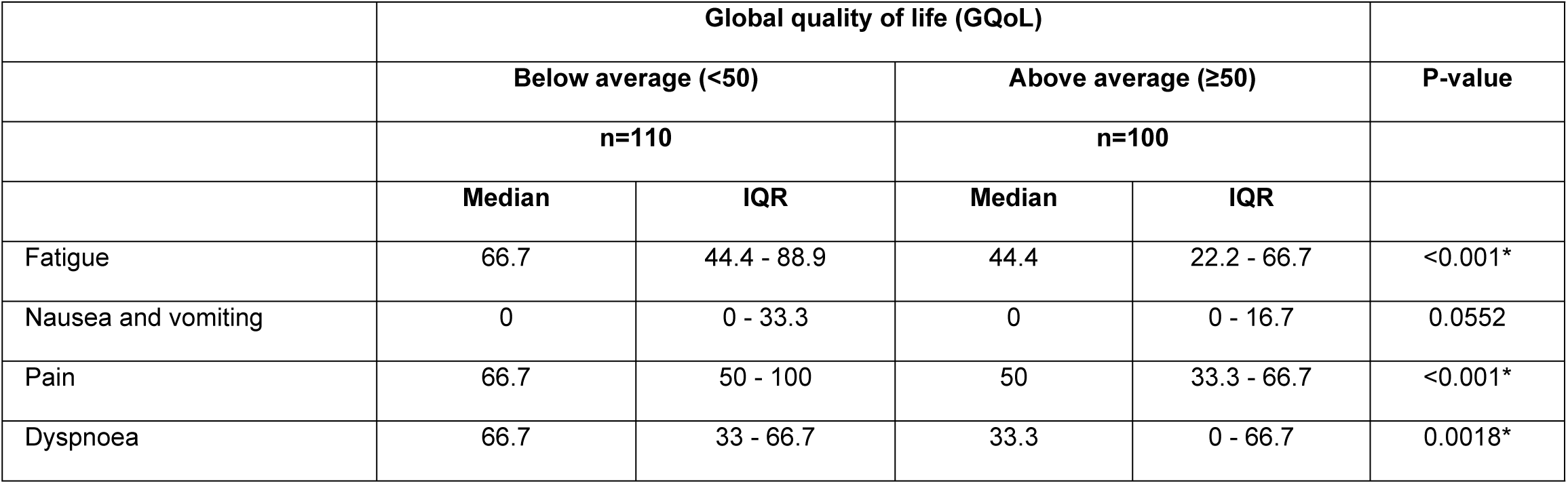
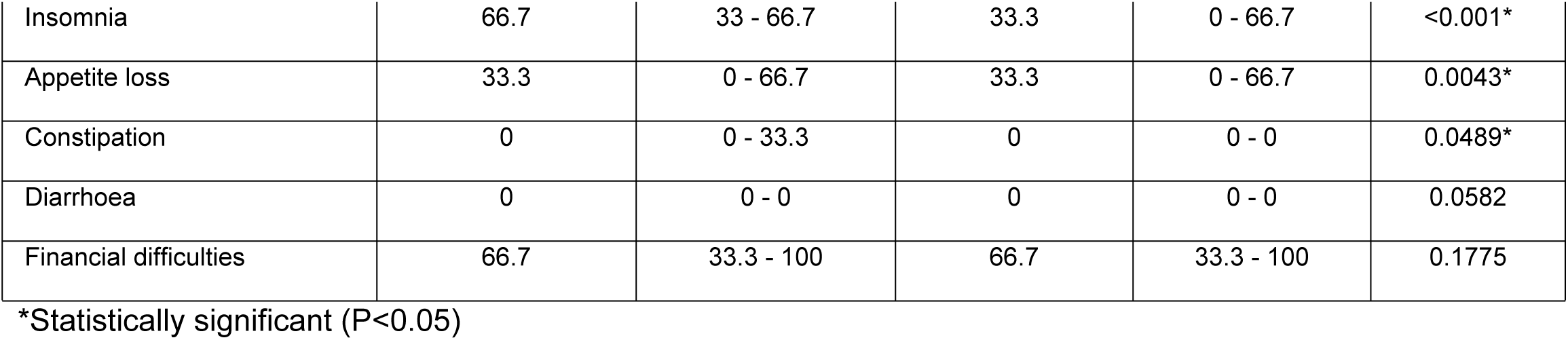
Associations between symptoms and financial difficulty with global quality of life.

|  | Global quality of life (GQoL) |  |  |  |  |
| --- | --- | --- | --- | --- | --- |
|  | Below average (<50) |  | Above average (≥50) |  | P-value |
|  | n=110 |  | n=100 |  |  |
|  | Median | IQR | Median | IQR |  |
| Fatigue | 66.7 | 44.4 - 88.9 | 44.4 | 22.2 - 66.7 | <0.001* |
| Nausea and vomiting | 0 | 0 - 33.3 | 0 | 0 - 16.7 | 0.0552 |
| Pain | 66.7 | 50 - 100 | 50 | 33.3 - 66.7 | <0.001* |
| Dyspnoea | 66.7 | 33 - 66.7 | 33.3 | 0 - 66.7 | 0.0018* |
| Insomnia | 66.7 | 33 - 66.7 | 33.3 | 0 - 66.7 | <0.001* |
| Appetite loss | 33.3 | 0 - 66.7 | 33.3 | 0 - 66.7 | 0.0043* |
| Constipation | 0 | 0 - 33.3 | 0 | 0 - 0 | 0.0489* |
| Diarrhoea | 0 | 0 - 0 | 0 | 0 - 0 | 0.0582 |
| Financial difficulties | 66.7 | 33.3 - 100 | 66.7 | 33.3 - 100 | 0.1775 |

\*Statistically significant (P<0.05)

## Discussion

This study aimed to describe the common symptoms, quality of life of lung cancer patients and association of demographic and clinical characteristics with global health status (GQoL). The EORTC GQoL for the lung cancer cohort in this study is low and differed across the three sites, with BMC and the Lung Lab reporting significantly lower GQoL compared to MTRH. The symptom burden is higher among patients with lower GQoL, with common symptoms being pain, fatigue, dyspnoea and insomnia, and appetite loss. There are significant differences in the socio-demographic, clinical characteristics, global health status, social functioning, common symptoms, and financial difficulties between the sites, as well as significant association between poor global health status, and ECOG performance and the type of symptoms.

### Quality of life

Our study has found that the GQOL of lung cancer patients at the three sites is low compared to the EORTC lung cancer reference sample (Median 58.3), consistent with previous studies^20^.Togas, in a Greek study of 200 lung cancer patients found that the GQoL and functionality among their cohort lower than the mean scores for the reference population, but slightly higher than that of our study: physical functioning (56.9), role functioning (48.9), and social functioning (50.5)^21^. Except for the emotional, all functional scales in our study are low compared to the reference sample. Poor physical and role functioning relates to daily activities that patients have to engage in, their need for assistance, ability to be productive at work and fulfilment of social roles. With advancing disease, most patients will require assistance with walking, washing, or bathing themselves and to fulfil their family responsibilities and roles as husband/wife, father/mother. Cognitive functioning among our participants is low, and also differed significantly between the sites, with MTRH the lowest at 58.3 (IQR 25-83.3)^11 22 23 24^. Cognitive functioning measures difficulty in remembering, concentration and paying attention. Previous studies have reported high cognitive functioning (82-89) among lung cancer patients compared to this study^11 22 23^. In a study of 139 lung cancer patients with stage III and IV, Yu Lee Dai reported cognitive functioning of 89.93 at baseline for lung cancer patients, concurring with previous reports^23^. The reason for the low cognitive functioning among the MTRH patients may be related to dyspnoea, pain and insomnia which are common among this cohort, and has been reported previously^25^. According to Chen among 115 lung cancer patients, insomnia negatively affected cognitive function. Cognitive ability reduces stress and symptoms and help patients cope^26^. Efforts to improve cognitive functioning may assist patients to cope with their disease.

### Sociodemographic and clinical characteristics

The study presents interesting differences in demographic and clinical characteristics between the three sites. The Lung Lab cohort differed from MTRH and BMC in that the participants were mostly male, had higher education levels, were likely to be smokers, had co-morbidities and likely to be diagnosed with stage 3 cancer, compared to stage 4 at other sites. Previous studies have reported that geographical distance can negatively affect access to cancer care^27^. In a qualitative study in KwaZulu Natal, Lubuzo highlighted distance from the facility as one barrier to accessing lung cancer care. The Lung Lab services a predominantly urban population compared to BMC and MTRH, which might explain the low number of stage 3 patients diagnosed at the two sites. Other reasons for the difference might be access to qualified radiologists available in South Africa. Further research to understand this difference is recommended.

Studies have previously reported association between patients’ sociodemographic characteristics and QoL. Research reports associating QoL with age, education, gender, economic status and GQoL are inconclusive, with some reporting better GQoL among older patients, males and those with higher level of education ^11^, while others reports better QoL among younger patients females ^28^. In a study 6,420 lung cancer patients in Texas, USA, Pierznski et al found that patients with higher level of education reported better GQoL^20 29^. Despite differences in sociodemographic characteristics between Wits-Core and the other sites in the current study, only the level of education was associated with GqoL. Literacy affects patients’ understanding of health information, motivation to seek help, application of information and making judgements^30^. The patients with the lowest level of education are at BMC and MTRH, both of which serve significant proportions of rural communities compared to the Lung Lab. Efforts to provide cancer health education in local languages and in rural communities in Kenya and Tanzania is recommended if we aim to improve early detection of lung cancer that will affect the QoL. Smoking is associated with poor quality of life and survival^31 25^. In a case control study of 168 cases and 334 controls, Chen et al (2015) found that smoking affected the quality of life as well as symptoms of lung cancer patients even after diagnosis^25^. We found no association between smoking and global health status GQoL, like what was reported by Togas et al^21^.

### Common symptoms

Advanced disease is associated with severe symptoms and poor QoL among lung cancer patients^11^. Almost all participants in our study had advanced disease. Common symptoms reported in this study include fatigue, pain, dyspnoea, insomnia, and loss of appetite, which is consistent with what has been reported elsewhere ^11 32 7 33^. The severity of pain and insomnia was higher amongst patients at BMC than at the Lung Lab and MTRH. Pain and insomnia have been reported among lung cancer patients elsewhere^7 21^. Higher pain scores among BMC may be related to access and availability and access of health services and pain medication. Challenges to the use of opioids for pain in SSA include strict laws controlling availability and prescription, health professional knowledge gaps and out of pocket expenditure among the poor communities^34^. To improve pain control in SSA, we advocate for policy change to allow for healthcare providers other than doctors to prescribe opioids for palliative care, make healthcare services affordable, and address the knowledge gaps among health professionals across all levels of care. Uganda is one of the countries that has allowed nurses trained in palliative care to prescribe morphine, a practice that can be emulated by other countries^35^. Fatigue, pain, dyspnoea, insomnia, and loss of appetite are associated with poor quality of life in our study. Similar findings have been reported by other studies^35^. Physical symptom management to improve quality of life is a core principle of palliative care, and is recognised as integral to good quality cancer care^37 38 39^. We recommend that all sites integrate palliative care as a component of cancer care to address the common symptoms and improve GQoL among lung cancer patients.

### Financial difficulties

Financial difficulty is associated with late presentation, poor GQoL, high symptom burden and non-adherence to treatment, but not in the current study^40 41 34^. Financial hardship may be direct (cost of health care services) or indirect (cost of transport, food, loss of income)^34 40 41^.

Most (80%) patients in our study were unemployed. Patients at BMC and MTRH reported greater financial difficulties than at Wits-Core or in other studies ^20 24^. A study of 150 cancer patients in Ethiopia, also found that cancer patients have financial difficulty^41^. Financial difficulty affects the ability to travel to health facilities for treatment, follow up visits, buying necessary food, supplements vitamins, and over the counter medicine^34 40 41^ . In the multi-site CanCORS study of 10,000 lung and colon cancer patients in the United States of America, Lathan reported a strong association between financial strain, poor QoL and high symptom burden^40^. The South African government provides social grants to patients disabled with severe limitations of activities of daily living including advanced cancer. We recommend that other countries in SSA consider allocating a budget towards similarly assisting patients with severe life-limiting illnesses to improve financial stability, thereby impacting on their quality of life.

Our study also has some limitations. The sample sizes at BMC and MTRH were small. While all sites used the standardised QLQ C30 tool, languages of interviews differed which may have affected the fidelity of data collection. Reported stages of lung cancer may have been affected by facilities available for accurate staging. Cough, which is one of the commonest symptoms of lung cancer is not included in the assessments and other unmeasured factors such as treatment effects, nutritional factors, use of alternative treatments such as herbal treatment and polypharmacy may have impacted on the GQoL. However, to our knowledge, this is the first study to describe the quality of life and common symptoms among patients diagnosed with lung cancer across three countries in SSA through collaborative research.

## Conclusion

This study found low levels of quality of life, physical functioning, role functioning and social functioning among lung cancer patients at three SSA sites, which was variable between sites. Poor QoL in the study is associated with level of education, performance status, fatigue, pain, dyspnoea, insomnia, loss of appetite and constipation. While all three sites had access to diagnostic tools, early diagnosis remains a challenge, especially in Kenya and Tanzania. There are no official screening programs for lung cancer in the three countries, due to constrained resources^42 43^. Risk factor modification, such as smoking cessation however has been at the forefront in cancer prevention. We recommend that more research be conducted to investigate the effectiveness of programs to reduce lung cancer risks, reasons for delays in diagnosis, the needs of lung cancer patients in different countries, and factors that may be impacting the quality of life. This research would guide interventions that best impact on the burden of lung cancer on patients in sub-Saharan Africa.

## Data Availability

The data used to support the research findings is included as part of the supporting information.

## Acknowledgements

The research was supported through grants from Bristol Myers Squibb Foundation. We acknowledge the support received from the clinical teams at Moi Teaching and referral Hospital (Kenya), Bugando Medical Centre (Tanzania) and Wits Centre for Palliative Care (South Africa).

